# COVision: Convolutional Neural Network for the Differentiation of COVID-19 from Common Pulmonary Conditions Using CT Scans

**DOI:** 10.1101/2023.01.22.23284880

**Authors:** Kush V. Parikh, Timothy J. Mathew

## Abstract

With the growing amount of COVID-19 cases, especially in developing countries with limited medical resources, it is essential to accurately and efficiently diagnose COVID-19. Due to characteristic ground-glass opacities (GGOs) and other types of lesions being present in both COVID-19 and other acute lung diseases, misdiagnosis occurs often — 26.6% of the time in manual interpretations of CT scans. Current deep-learning models can identify COVID-19 but cannot distinguish it from other common lung diseases like bacterial pneumonia. Concretely, COVision is a deep-learning model that can differentiate COVID-19 from other common lung diseases, with high specificity using CT scans and other clinical factors. COVision was designed to minimize overfitting and complexity by decreasing the number of hidden layers and trainable parameters while still achieving superior performance. Our model consists of two parts: the CNN which analyzes CT scans and the CFNN (clinical factors neural network) which analyzes clinical factors such as age, gender, etc. Using federated averaging, we ensembled our CNN with the CFNN to create a comprehensive diagnostic tool. After training, our CNN achieved an accuracy of 95.8% and our CFNN achieved an accuracy of 88.75% on a validation set. We found a statistical significance that COVision performs better than three independent radiologists with at least 10 years of experience, especially in differentiating COVID-19 from pneumonia. We analyzed our CNN’s activation maps through Grad-CAMs and found that lesions in COVID-19 presented peripherally, closer to the pleura, whereas pneumonia lesions presented centrally.

## 1 Introduction

### 1.1 Background

The outbreak of severe acute respiratory syndrome coronavirus-2 (SARS-CoV-2) and its associated disease COVID-19 has led to a global pandemic. As of March 31, 2022, there have been over 486 million COVID-19 cases worldwide, claiming an estimated 6.14 million lives according to the World Health Organization (WHO) [1]. COVID-19 infects the lungs, specifically the alveolar type II cells, resulting in complications like pneumonia [2]. Currently, RT-PCR remains the gold standard for COVID-19 diagnosis; however, due to limited sensitivity of 89.9% [3], and a wait time of at least 48 hours for results, the need for quicker and more accurate diagnosis is imperative. This is especially the case when patients present to the hospital with severe respiratory diseases that could be COVID-19 or other conditions with similar presentations such as bacterial pneumonia, pulmonary edema, or sepsis. Because of the similarity in the presentation of these pulmonary conditions, it is often difficult to form an accurate diagnosis with CT scans alone, leading to a high rate of misdiagnosis.

### 1.2 Disproportionate Effect of COVID-19

The disparity in the COVID-19 response between developing and developed countries is staggering. According to the World Bank, high and high-intermediate countries have higher physicians per capita and nurses per capita when compared to low and low-intermediate-income countries [4]. Factors such as slow economic growth in developing countries and the migration of healthcare workers from developing to developed countries are the primary reasons attributed to the lack of healthcare professionals in developing nations. The shortage of healthcare workers in low and low-intermediate countries has led to greater work hours per week and higher rates of burnout [5]. These issues have only been exacerbated due to the COVID-19 pandemic leading to overburdened medical systems. Using digital technology and automation in healthcare, particularly in low-income nations, has great potential to ease the burden on their already crumbling medical infrastructure.

### 1.3 Deep Learning

New developments in deep learning have led to potential diagnostic applications. Deep learning allows for the extraction of nonlinear quantitative features in datasets allowing for analysis of complex patterns in the training data, leading to the possibility of creating automated high-accuracy diagnosis models in radionomics [6]. In the past, convolutional neural networks (CNNs) have shown general usability in diagnosing retinal conditions using optical coherence tomography [7].

### 1.4 Existing Work

SARS-Net [8] is one of many deep-learning models developed to aid with COVID-19 diagnosis. While this model is able to achieve an accuracy of 97.6% in identifying COVID-19 from Chest X-rays (CXRs), this model fails to differentiate COVID-19 from other common pulmonary conditions such as bacterial pneumonia leading to a low specificity. Specificity is a measure of how well a model can identify individuals who do not have a disease and can correctly identify what condition(s) an individual might have instead. For effective use in a clinical setting, and for triaging of patients, models that detect COVID-19 from medical images and CT scans need a high specificity. One potential solution is to apply Capsule Networks [9]. However, these networks use vector representation for the neurons and replace the backpropagation training algorithm with dynamic routing, both of which substantially increase computational complexity and training time.

### 1.5 Introduction

In our research, we attempted to answer the following research question: Is it possible to create a lightweight computer model that can accurately identify COVID-19 and distinguish it from other common lung diseases, such as bacterial pneumonia? Due to the aforementioned potential of deep learning, we hypothesize that we can create a lightweight model, called COVision, using deep learning that will be on par or outperform radiologists in the differentiation of COVID-19 and bacterial pneumonia as measured by the metrics of accuracy, AUROC, and sensitivity (recall), specificity, and precision.

Our significant contributions are listed below:

- We developed a lightweight model called COVision that can accurately diagnose and differentiate COVID-19, bacterial pneumonia, and healthy patients using CT scans and basic clinical factors.
- Our model outperformed three independent radiologists on a testing set with statistical significance.
- We made the following observations: Upon analysis of the Grad-CAMs for our CNN, we found that lesions in COVID-19 presented peripherally, closer to the pleura, whereas pneumonia lesions presented centrally. Additionally, “shortness of breath” is the most influential clinical factor for diagnosis.

## 2 Methods

### 2.1 CT Scans Preprocessing

194,922 isolated CT slices (75,686 COVID-19, 73,924 pneumonia, 45,312 healthy) were obtained from the CC-CCII dataset [10]. All 194,922 data points are anonymized and deindividualized. These CT slices were split 80/20 into training and testing sets (60,548/15,138 COVID-19, 59,139/14,785 pneumonia, 36,249/9,063 healthy). To standardize the images, they were resized to 512 × 512 x 1 through Lanczos3 interpolation. Layers of augmentation such as alterations to brightness, saturation, and rotations were then applied to training images. The complete flow of data is shown in Table 1.

**Table 1:** Breakdown of how many CT scans and clinical factors were used for the testing and training.

|  | Condition | Original Dataset |  | Sample Data |  |
| --- | --- | --- | --- | --- | --- |
|  |  | Training | Testing | Training | Testing |
| CT Scans | COVID-19 | 60548 | 15138 | 35000 | 5638 |
|  | Pneumonia | 59139 | 14785 | 35000 | 7254 |
|  | Healthy | 36249 | 9063 | 35000 | 12766 |
| Clinical Factors | COVID-19 | 9600 | 2400 | 9600 | 2400 |
|  | Pneumonia | 9600 | 2400 | 9600 | 2400 |
|  | Healthy | 9600 | 2400 | 9600 | 2400 |

### 2.2 Proposed Convolutional Neural Network

We used the Keras API in Tensorflow to design, train, and test our CNN. The input for our CNN is a processed image, and the output is a probability distribution of predicted classes (COVID-19, bacterial pneumonia, and healthy). The specific architecture for our model is shown in Figure 1. In our model, we used maximum pooling to reduce the size of feature maps. Brighter pixels have grayscale values closer to 1 while darker pixels have grayscale values closer to 0. This is why maximum pooling over other options such as minimum pooling because on CT scans the maximum values (i.e. the brightest pixels) contain the most relevant information about the image needed for the classification and diagnosis of lung diseases. Specifically, common lesions in COVID-19 and bacterial pneumonia such as GGOs, crazy-paving patterns, and consolidation in the lungs all show up on a CT scan as brighter pixels. Our CNN has 6,542,531 trainable parameters.

**Figure 1:**
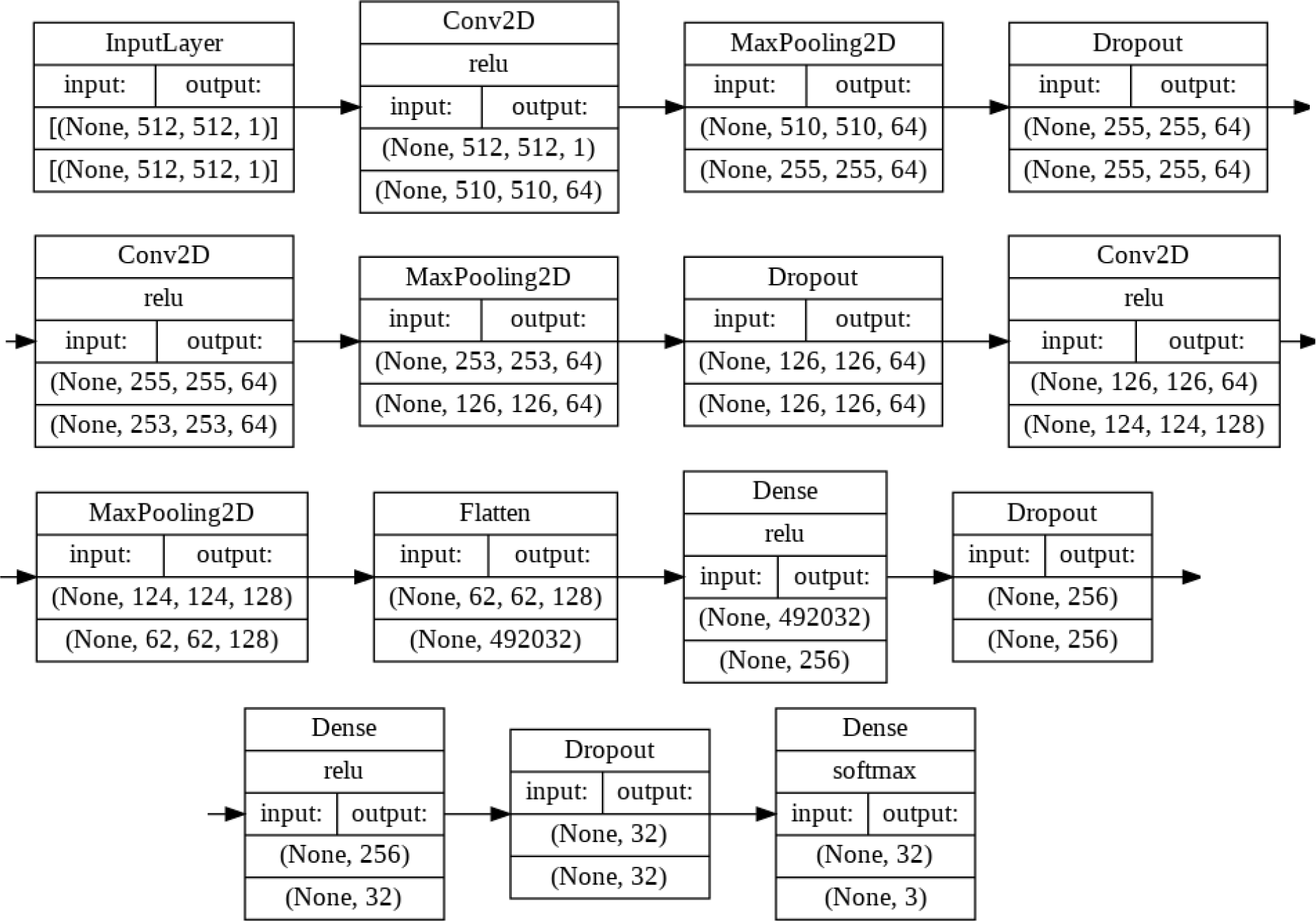
Visualization of our CNN’s architecture. The diagram was displayed by using Tensorflow in Python.

#### 2.2.1 Minimizing Complexity

Current state-of-the-art models such as VGG19, InceptionV3, and ResNet152 have 19, 48, and 152 layers respectively. The training accuracy of a CNN will generally increase with more hidden layers, along with the computational complexity of the model. However, an overly complex model will often overfit the training data and underperform on the testing data because it learns the patterns in the training data so well that it is not able to extrapolate to testing data. Another issue is that on computing systems with low computation power (i.e. simple desktop CPUs [11]) as is the case with many hospitals, these models may be untrainable due to the immense amount of computation required. In medical imaging there typically is a tradeoff between the efficiency and the accuracy the model achieves on testing data [12]. Our CNN is a lightweight model with only 6 hidden layers. Yet, as we demonstrate it is able to achieve superior performance as shown in Section 3.1.

#### 2.2.2 Dropout Layers

We further prevented overfitting by implementing regularization through dropout. We placed dropout layers after the 1st and 2nd max-pool layers and after the 1st and 2nd dense layers (Figure 1). The standard convention is to set dropout p = 0.5 for fully connected (dense) layers and p = 0.8 or 0.9 for convolutional layers, however, this technique is arbitrary and is not generalizable to every model. Using GridSearchCV from the sklearn library, we use grid searching to test dropout factors between 0.1 and 0.9 (increment of 0.1) in combination for all four dropout layers. The following set of dropout factors achieved the highest accuracy on a training set: 0.6 for both the convolutional layers and 0.7 for both the dense layers.

### 2.3 Training Convolutional Neural Network

Our CNN was trained using a stratified random sample of 105,000 isolated CT slices taken from our training set created in Section 2.1 (Table 1). We used 35,000 slices each for COVID-19, pneumonia, and healthy (control). We initialized the weights for our model using the Glorot (Xavier) Uniform Initializer. We used categorical cross-entropy as our loss function. Adam’s optimizer was used to minimize the loss. The hyperparameters for Adam’s optimizer were tuned using a grid-search method. The combination that achieved the lowest root mean squared error (RMSE) is summarized in Table 2. We trained our CNN on an NVIDIA GeForce 3090 GPU for 250 epochs (Figure 2). Scripts were run on Ubuntu 20.04.

**Table 2:**
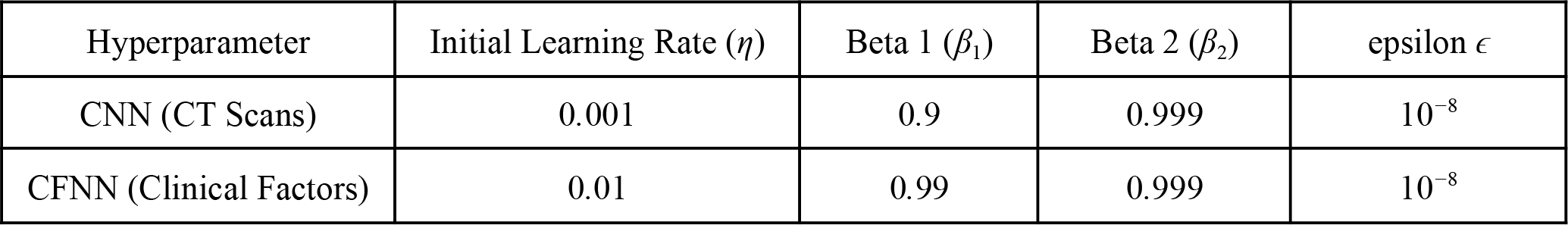
CNN and CFNN Adam’s optimizer hyperparameters that achieved lowest RMSE after grid-searching.

**Figure 2:**
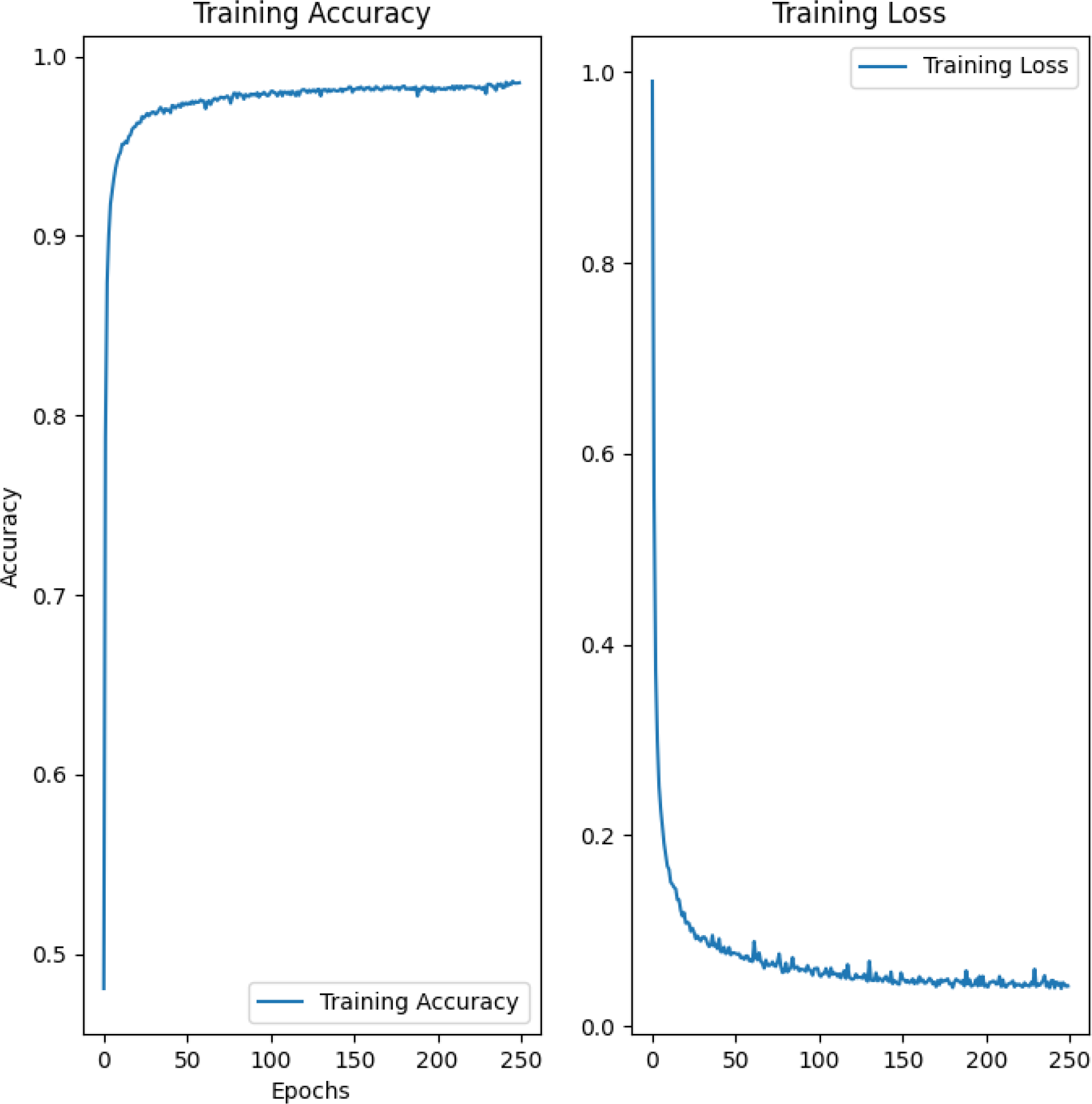
Accuracy (left) and loss (right) of the CNN in classifying the training data across 250 epochs.

### 2.4 Clinical Factors Preprocessing

We used the Khorshid COVID Cohort (KCC) [13] and the Israeli Ministry of Health public health database [14] to construct a custom dataset of 7 clinical factors (shortness of breath, cough, headache, fever, sore throat, age, and gender). Factors such as shortness of breath, cough, headache, fever, sore throat, and gender were expressed as a boolean. Age was expressed as an integer. In total, there were clinical factors for 30 COVID-19 patients, 30 bacterial pneumonia patients, and 125,882 healthy patients. All 125,942 data points came from unique, anonymized, deindividualized patients.

Training a model on this dataset results in an imbalance classification problem because of the skewness in distribution over the three classes. To address this, the data was resampled using the Imbalanced Learn library in Python. The majority class of healthy patients was sampled so that 12,000 sets of clinical factors were randomly selected. Both minority classes of patients with COVID-19 and patients with pneumonia were oversampled through random duplication so 11,970 sets of clinical factors were added to the original 30 sets for both classes. Specifically, we used the synthetic minority oversampling technique (SMOTe) which is a sampling strategy of 1:399 for the COVID-19 and the pneumonia classes. The complete dataset had 36,000 data points each with sets of clinical factors equally distributed among the three classes. These were split 80/20 into training and testing sets (9,600/2,400 COVID-19, 9,600/2,400 pneumonia, 9,600/2,400 healthy).

### 2.5 Clinical Factors Neural Network (CFNN)

In addition to CT scans, a patient’s clinical factors can serve as a means of differentiating whether a patient has COVID-19 or bacterial pneumonia. We designed this neural network called the clinical factors neural network (CFNN) to work in conjunction with our CNN (for CT Scans) that was described in Sections 2.2 and 2.3 While a CNN uses convolutional layers to classify CT scans, the CFNN is a standard multilayer perceptron neural network used to classify sets of quantitative factors such as age. The ensembling process to combine the CFNN and CNN is described in Section 4.

We used the Keras API in Tensorflow to design, train, and test our CFNN. The input for our CFNN is a processed set of clinical factors, and the output is a probability distribution of predicted classes (COVID-19, bacterial pneumonia, and healthy). We chose our CFNN to have 6 fully connected layers. The specific architecture for our model is shown in Figure 3. We added a dropout layer for regularization after the first 3 dense layers to reduce overfitting. The dropout factor was tuned to p = 0.5 using the same grid-searching method in Section 2.2.2. There are 60,099 trainable parameters.

**Figure 3:**
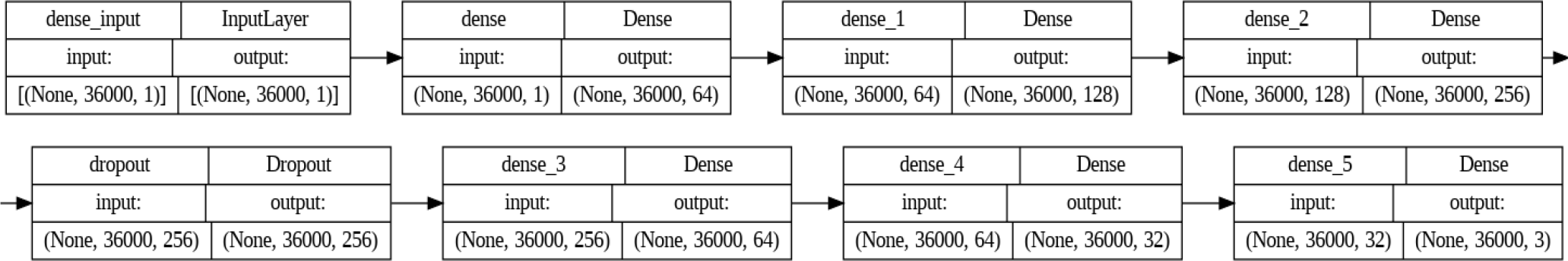
Visualization of our CFNN’s architecture. The diagram was displayed by using Tensorflow in Python.

### 2.6 Training CFNN

We trained with 28,800 sets of clinical factors (9,600 COVID-19, 9,600 bacterial pneumonia, and 9,600 healthy). The accuracy and loss of the model began to stabilize by 40 epochs so we early stopped training the model at 50 epochs (Figure 4). The weights of our CFNN were initialized using a Glorot Uniform Initializer. We used categorical cross-entropy as our loss function. Adam’s Optimizer was used to optimize the weights. We used grid searching to choose the optimal hyperparameters (Table 2). We trained our CFNN on an NVIDIA GeForce 3090 GPU. Scripts were run on Ubuntu 20.04.

**Figure 4:**
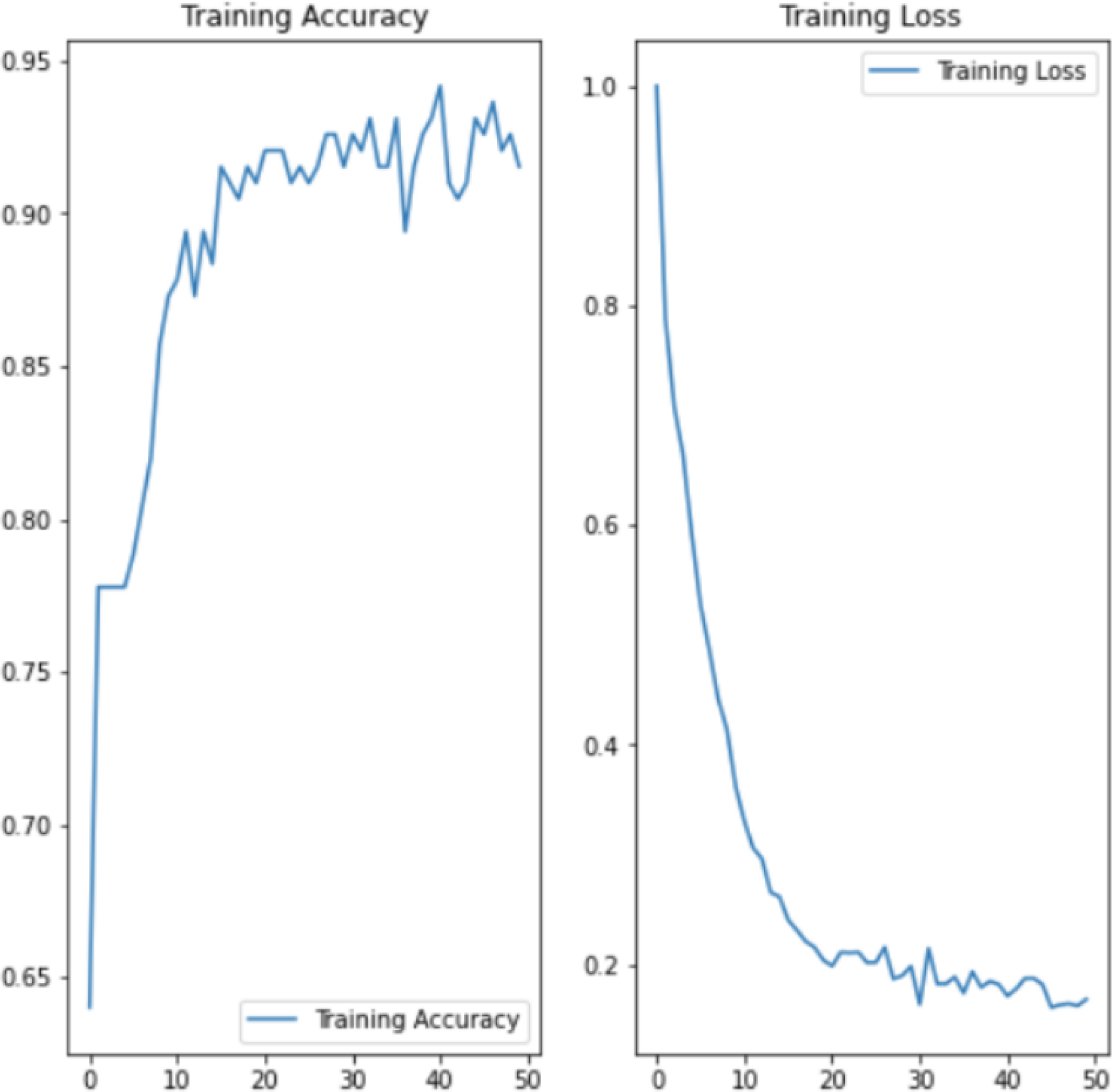
Accuracy (left) and loss (right) of the CFNN in classifying the training data across 50 epochs.

## 3 Results

### 3.1 CNN Testing

To test our CNN, we took a stratified random sample of 25,658 CT slices from our testing set created in Section 2.1 (Table 1). The breakdown of the testing data is as follows: 5,638 COVID-19, 7,254 pneumonia, and 12,766 healthy (control). The confusion matrix of the results after classifying the testing images using our CNN is summarized in Figure 5. The accuracy, AUROC, sensitivity (recall), specificity, and precision of the CNN on the testing images are summarized in Table 3. For this classification problem, we calculated the overall AUROC to be 0.970 using weighted averaging of the class AUROCs.

**Table 3:** Accuracy, AUROC, sensitivity (recall), specificity, and precision of the CNN compared to radiologists.

| Metric | COVision |  |  | Radiologists |  |  |
| --- | --- | --- | --- | --- | --- | --- |
|  | Healthy (control) | Pneumonia | COVID | Healthy (control) | Pneumonia | COVID |
| Accuracy | 0.970 | 0.981 | 0.967 | 0.966 | 0.758 | 0.744 |
| AUROC | 0.982 | 0.967 | 0.961 | 0.976 | 0.793 | 0.766 |
| Sensitivity (Recall) | 0.949 | 0.976 | 0.956 | 0.985 | 0.356 | 0.439 |
| Specificity | 0.990 | 0.983 | 0.969 | 0.951 | 0.964 | 0.934 |
| Precision | 0.989 | 0.957 | 0.896 | 0.942 | 0.837 | 0.807 |

**Figure 5:**
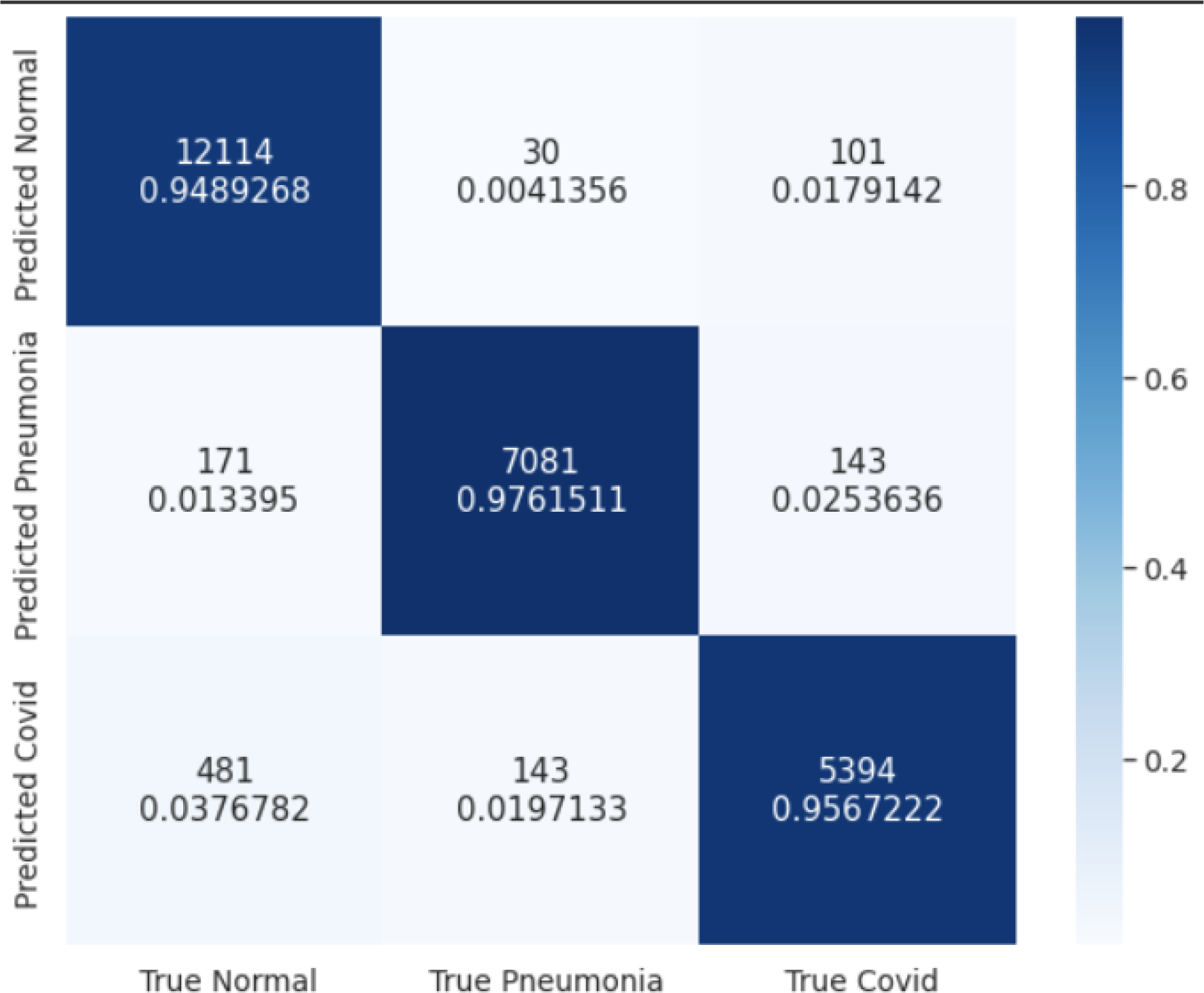
Confusion matrix comparing the true labels for the images and the predicted labels by our CNN.

For COVID-19 detection, a high sensitivity (recall) measures that our model minimizes the number of positive cases that are missed, which would lead to further spread of the virus if not addressed quickly. A high specificity indicates that the model effectively minimizes the number of healthy individuals who are incorrectly classified as COVID-19 positive, which could save the healthcare system unnecessary costs of treatment and care and prevent unneeded quarantine. Similar to high specificity, high precision ensures that the amount of true diagnoses is maximized, increasing efficiency within the hospital.

### 3.2 Comparison Against Radiologists

We emailed radiologists with at least 10 years of experience from Ascension Hospital, Henry Ford Hospital, and Beaumont Hospital to participate in our study. We took a random sample of 297 images from our testing set and asked three radiologists to classify CT scans as either COVID-19, bacterial pneumonia, or healthy. Radiologist 1 classified 97 images, Radiologist 2 classified 150 images, and Radiologist 3 classified 88 images. The radiologists’ combined results are summarized in Figure 6. Our model had an accuracy of 95.8% while the radiologists had an accuracy of 73.4% (p < 0.0001). When analyzing the confusion matrices (Figures 5 and 6), we find that our CNN can differentiate COVID-19 from bacterial pneumonia with 97.8% accuracy while the three trained radiologists differentiate with a 55.5% accuracy. A full comparison of our CNN’s accuracy, AUROC, sensitivity (recall), specificity, and precision when compared to the radiologists can be found in Table 3.

**Figure 6:**
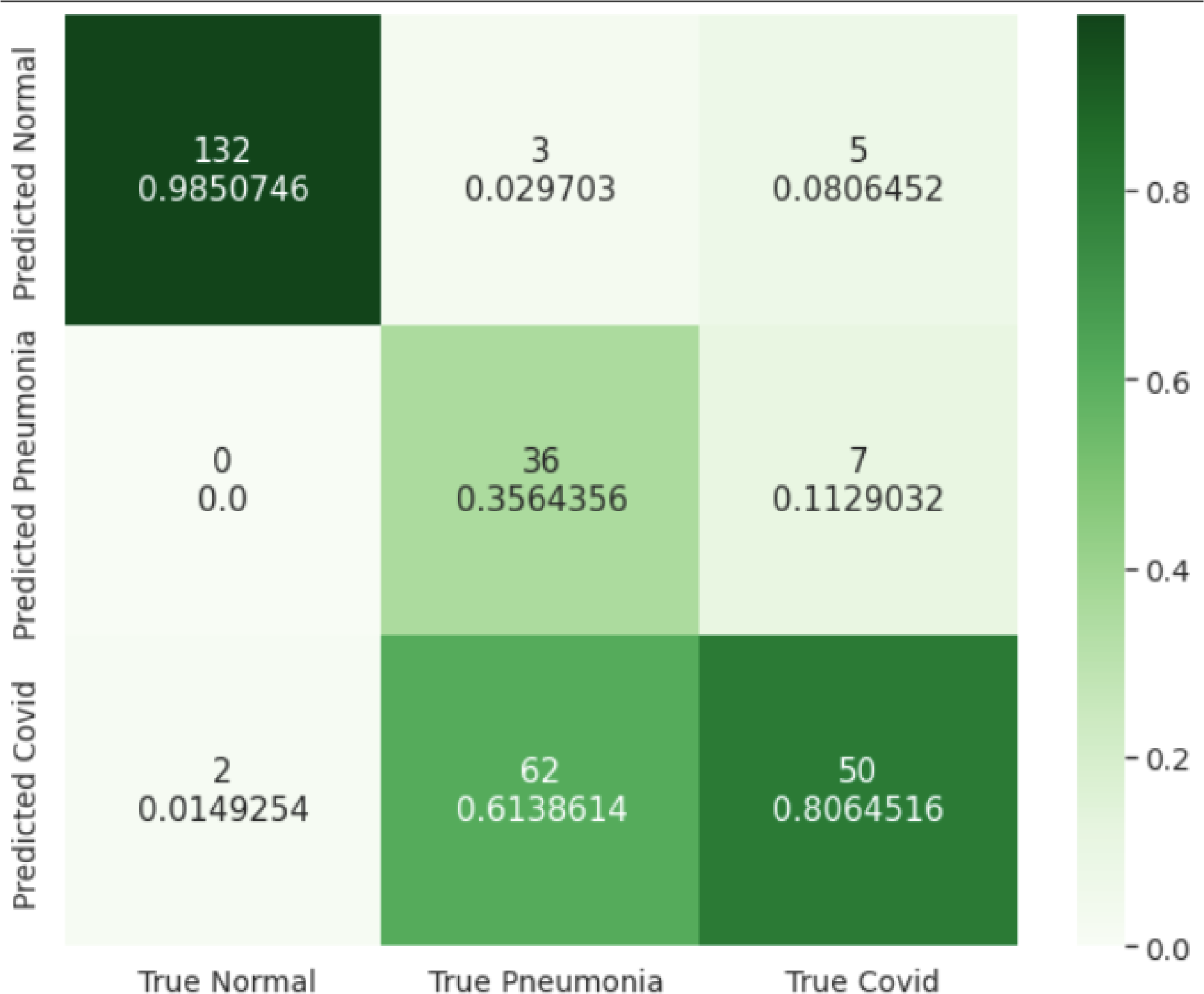
Confusion matrix comparing the true labels for the images and the predicted labels by the radiologists.

### 3.3 Grad-CAMs

To visualize the weights of the trained CNN, we created Gradient-Weighted Class Activation Mapping (Grad-CAMs) [15] for a stratified random sample of 2,000 CT slices (1,000 bacterial pneumonia, and 1,000 COVID-19) from our CT scan testing set without any data augmentation (i.e. flips, rotations, etc.) because we wanted to generalize our Grad-CAMs to a standard view of Chest CT Scans. Heat maps of the activation map from the CNN’s last convolutional layer were created with a CT scan as input. This quantitative heat map was then normalized to a range of [0, 1], averaged across all images in the class, and transformed into a single visualization using the *Matplotlib* library (Figure 7). The Grad-CAMS shows that lesions are generally present in the center of the lungs in bacterial pneumonia. Lesions for COVID-19 typically present peripherally, closer to the pleura. COVID-19 lesions are also shown to be much more scattered while lesions from bacterial pneumonia are more localized. These image features can be used by radiologists to improve the accuracy of manual diagnosis of conditions.

**Figure 7:**
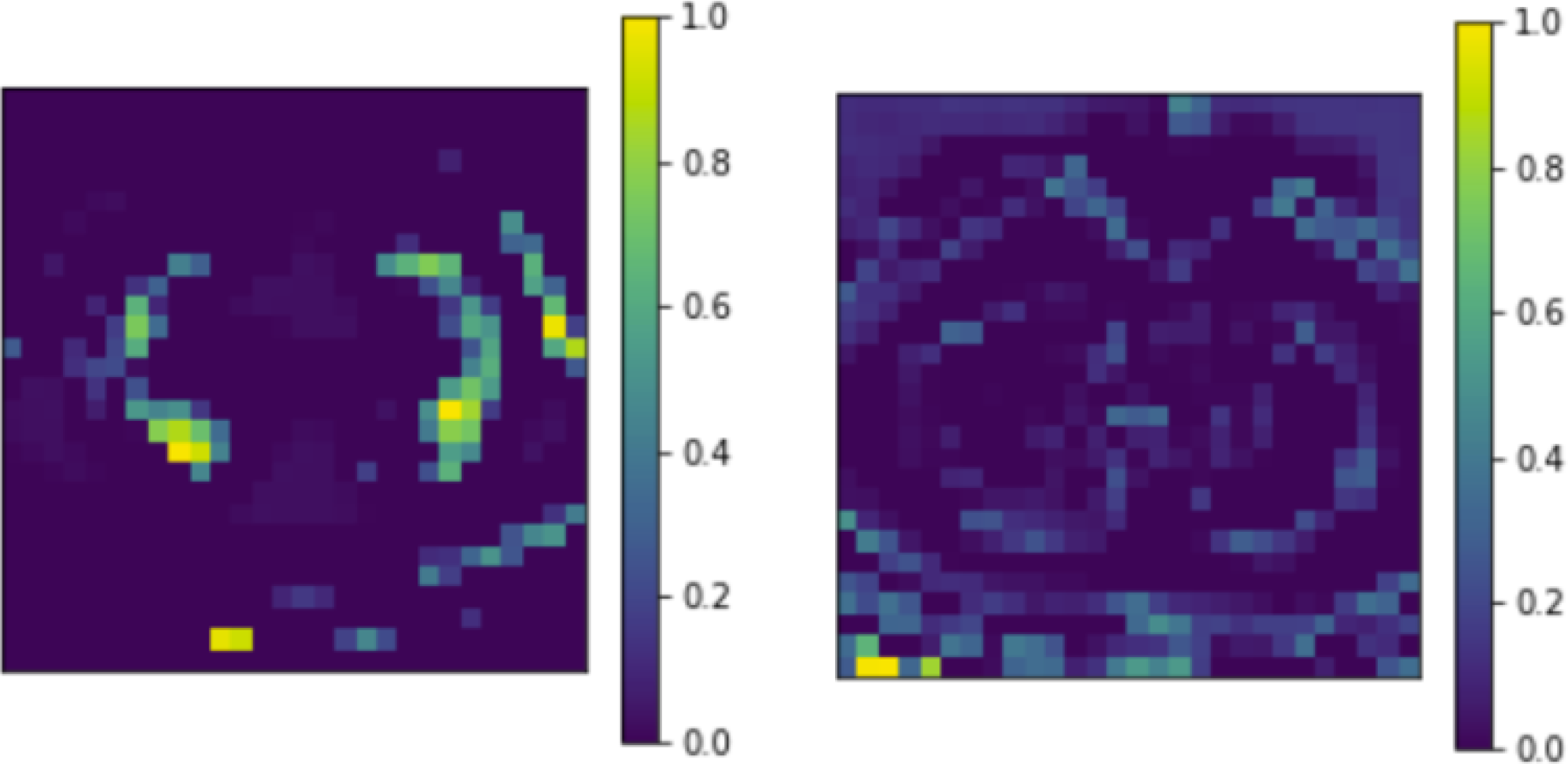
Grad-CAMs (average of 1000 images) for bacterial pneumonia (left), and COVID-19 CT scans (right).

### 3.4 CFNN Testing

To test our CNN, we took a stratified random sample of 7,200 sets of clinical factors from our testing set created in Section 2.1 (Table 1). The breakdown of the testing data is as follows: 2,400 COVID-19, 2,400 bacterial pneumonia, and 2,400 healthy (control) The CFNN achieved an accuracy of 88.75%. The confusion matrix of the results after classifying the patients using their clinical factors using our CFNN is summarized in Figure 8. The highest categorical accuracy of 97.58% came from the healthy class, however, this was followed by 85.46% for COVID-19 and 83.20% for bacterial pneumonia. Because of this, we realized that our CFNN should be used in conjunction with other models to produce the most accurate diagnosis. Because of this, we propose an ensemble model combining our CNN and CFNN in Section 4.

**Figure 8:**
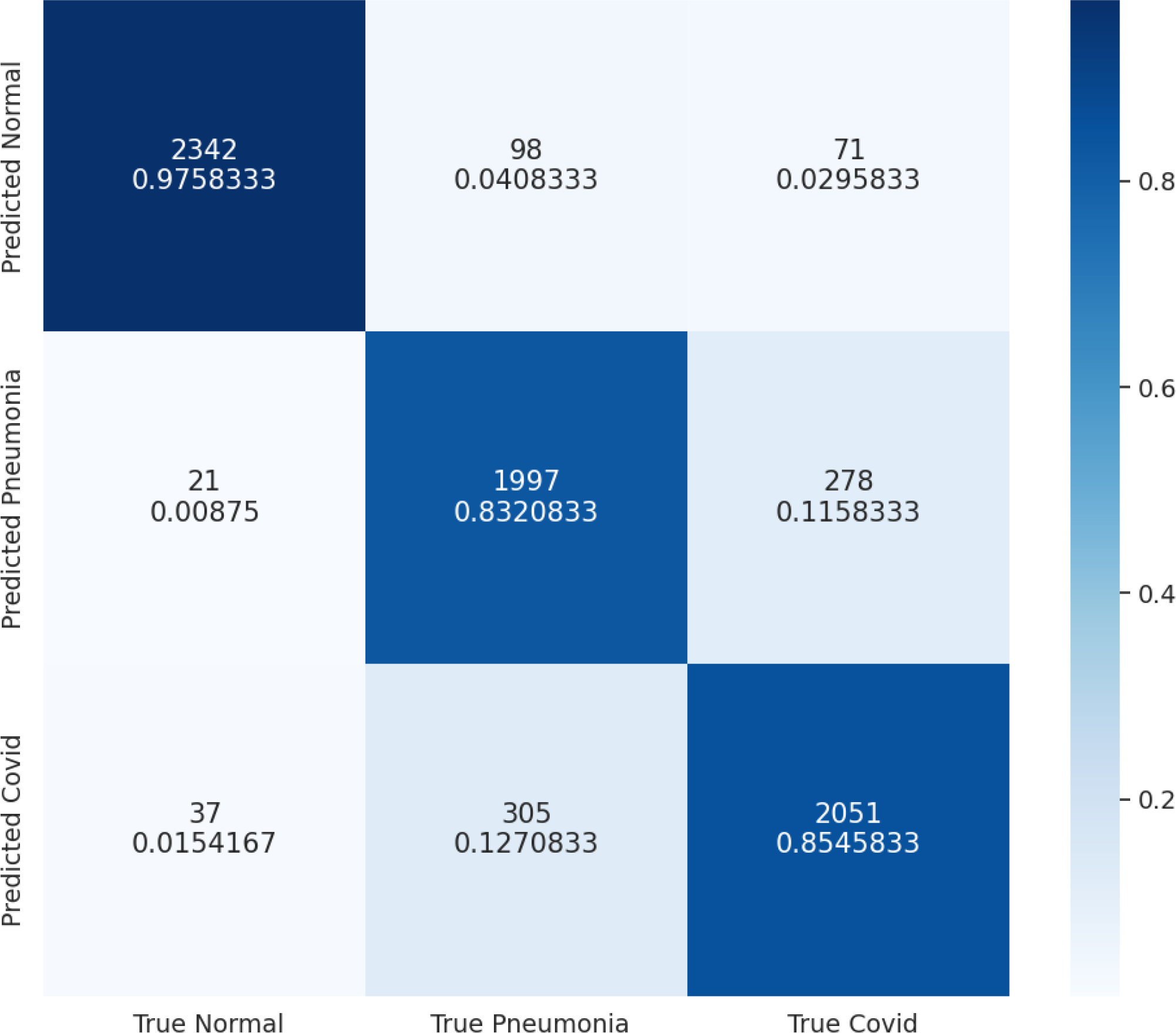
Confusion matrix comparing the true labels for clinical factors and the predicted labels by our CFNN.

### 3.5 CFNN Weights

The weights from the trained CFNN were extracted to determine the importance of each clinical factor in making a prediction. After normalizing the weights, we found that the most influential factor was “shortness of breath”.

## 4 Ensembling

To combine our CNN and CFNN which can both independently differentiate between healthy, bacterial pneumonia, and COVID-19 patients, we create an ensemble model. Specifically, the predictions of each network are combined using federated weight averaging [16]. The weight is based on the ratio of training data used for both models. Because the CNN was trained on 105,000 images and the CFNN was trained on 28,800 sets of clinical factors, the ratio is 0.785 to 0.215. Concretely, the prediction of the CNN has a weight of 0.785 while the prediction of the CFNN has a weight of 0.215.

## 5 Discussion

Through our research, we developed a deep learning framework to differentiate COVID-19 from other common pulmonary conditions such as bacterial pneumonia. Our framework has two parts: a convolutional neural network (CNN) that uses CT scans, and a clinical factors neural network (CFNN) that uses clinical factors such as age, gender, and symptoms to help diagnose and differentiate between healthy, bacterial pneumonia, and COVID-19 patients. Together we call this framework COVision. When compared to three board-certified radiologists with at least 10 years of experience, our CNN has a statistically significant higher accuracy of 95.8% to 73.4% (p < 0.0001), especially in differentiating COVID-19 from pneumonia and healthy CT Scans. Our CFNN on the other hand achieved an accuracy of 88.75% on the testing set. Particularly, our CFNN performs poorly at differentiating COVID-19 from bacterial pneumonia which is why we suggest the use of a CNN and CFNN ensemble model for a more comprehensive diagnosis. We also made a few observations that may be relevant for clinical practice. After constructing Grad-CAMs for our CNN we found that COVID-19 lesions presented peripherally, closer to the pleura while pneumonia lesions presented centrally on a chest CT scan of the lungs. After analyzing the weights of our trained CFNN, we found that “shortness of breath” was the best indicator for disease. Our research has two limitations. 1) Our data for the CT scans and for the clinical factors came from different patients which makes the ensembling process not as robust as if they came from the same patients. 2) The data for the CT scans and clinical factors came from patients from different parts of the world (i.e East Asia and Middle East). This could be an issue if COVID-19 and bacterial pneumonia present in the lungs differently in different parts of the world. In future, both of these problems can be ameliorated as more public patient data becomes available. In the future, with more data, the COVision can also be trained to differentiate other lung conditions apart from bacterial pneumonia such as different types of lung cancer. COVision has the potential to save countless lives, particularly in developing nations with a shortage of doctors and a huge volume of patients due to the coronavirus pandemic by assisting medical professionals in the diagnosis process.

## Data Availability

All data produced are available online at:
1) Consortium of Chest CT Image Investigation (CC-CCII) Dataset: http://ncov-ai.big.ac.cn/download?lang=en
2) Khorshid COVID Cohort (KCC): https://doi.org/10.6084/m9.figshare.16682422.v1
3) Israeli Ministry of Health: https://data.gov.il/dataset/covid-19/resource/74216e15-f740-4709-adb7-a6fb0955a048

https://doi.org/10.6084/m9.figshare.16682422.v1

https://data.gov.il/dataset/covid-19/resource/74216e15-f740-4709-adb7-a6fb0955a048

## 6 Declarations

### Ethics Approval and Consent to Participate

The original study from which the CT scans of COVID-19, bacterial pneumonia, and healthy patients were analyzed states that Institutional Review Board approvals were obtained from Sun Yat-sen Memorial Hospital and Third Affiliated Hospital of Sun Yat-sen University, The First Affiliated Hospital of Anhui Medical University, West China Hospital, Nanjing Renmin Hospital, Yichang Central People’s Hospital, and the Renmin Hospital of Wuhan University. The study specifies that the work was conducted in accordance with the Chinese CDC policy on reportable infectious diseases and the Chinese Health and Quarantine Law, and was adherent to the tenets of the Declaration of Helsinki. The original study from which the clinical factors for COVID-19 and pneumonia patients were analyzed states that the study was approved by the ethics committee of the Isfahan University of Medical Sciences (IUMS). The database from which the clinical factors for healthy patients were analyzed declares that all data was collected and held in accordance with the policies of the Privacy Protection Authority and with the guidelines of the Government Information and Communications Technology (ICT) Authority directed by the Israel National Digital Agency. All methods were carried out in accordance with BioMed Central guidelines. Written informed consent was obtained from all the subjects whose CT scans or clinical factors were analyzed.

### Consent for Publication

Not applicable.

### Availability of Data and Materials

The dataset for the CT scans of COVID-19, bacterial pneumonia, and healthy patients is available in the China Consortium of Chest CT Image Investigation (CC-CCII) [http://ncovai.big.ac.cn/download?lang=en]. The dataset for the clinical factors for COVID-19 and bacterial pneumonia patients is available in the Khorshid COVID Cohort (KCC) [https://figshare.com/articles/dataset/COVID-19_and_non-COVID-19_pneumonia_Dataset/16682422]. The dataset for the clinical factors for healthy patients is available from the Israeli Ministry of Health [https://data.gov.il/dataset/covid-19]. Ground truth for COVID-19 patients was established via a confirmed reverse-transcriptase-PCR. Ground truth for bacterial pneumonia was established via standard clinical, radiological, and culture/molecular assay results. CT scans were screened for quality control by removing unreadable scans.

The code used for our research is available on GitHub at: https://github.com/Kushy0814/COVision/tree/main.

### Authors’ Contributions

All authors contributed equally to this work.

## Acknowledgments

Not applicable.

